# Mode of Birth and Adherence to WHO Physical Activity Guidelines: A Cross-Sectional Study of Danish Postpartum Mothers

**DOI:** 10.1101/2025.11.28.25341203

**Authors:** Marie Graae Dammand, Solvej Videbæk Bueno, Sebastian Dyrup Skejø

## Abstract

**Aim:** The objective of this paper is to investigate if the prevalence proportion ratio of non-adherence to the World Health Organisation (WHO) guidelines of at least 150 minutes of moderate-to-vigorous physical activity (MVPA) per week differed among Danish mothers based on mode of birth. We hypothesised that women who gave birth via caesarean section adhere less to the WHO’s recommendations compared with women who gave birth vaginally.

**Method:** This population-based cross-sectional study is based on data from 1,919 Danish mothers participating in the Danish National Health Survey from 2021 and the Danish Medical Birth Register. Mode of birth was dichotomised into either caesarean section or vaginal birth based on diagnostic codes in the Danish Medical Birth Register. Physical activity measured as weekly minutes of MVPA based on the self-reported physical activity in the Danish National Health Survey. Based on the answers of the survey, the mothers were dichotomised into two groups: adherers to the WHO guidelines of PA and non-adherers to the WHO guidelines of PA. The prevalence proportion of non-adherence to WHO guidelines was compared between the two modes of birth.

**Findings:** The prevalence proportion of non-adherence was 2% lower among mothers who gave birth via caesarean section compared with mothers who gave birth vaginally, but with a 95% confidence interval ([0.90; 1.06]) indicating a statistically non-significant difference.

**Conclusion:** This study found no statistically significant difference in non-adherence to the WHO PA guidelines between mothers who gave birth via caesarean section and mothers who gave birth vaginally.

## Introduction

Physical activity (PA) is suggested to have numerous health benefits. Among these are reduced risk of developing cardiovascular disease, hypertension, specific types of cancer and type-2 diabetes[1]. Besides these physical benefits, PA also has many mental health benefits including an increase in well-being and a reduction in depressive and anxiety symptoms[2, 3]. Other mental health benefits of physical activity are improved self-concept and body image[4].

The World Health Organisation (WHO) recommends that adults (18-64 years) do at least 150 minutes of moderate-intensity PA or at least 75 minutes of vigorous-intensity PA per week or an equivalent combination of the two[5]. Nonetheless, the prevalence of insufficient PA is increasing, with a global age-standardised prevalence rising from 26.4% in 2010 to 31.3% in 2022[5, 6].

To combat insufficient PA in the population, the WHO suggests that nations implement national PA guidelines, while underlining the importance of the guidelines being disseminated to specific subgroups of people in a population[5]. One such subgroup could be mothers. Danish mothers tend to be less compliant with the WHO guidelines than nulliparous Danish women, with the proportion of parous women non-adhering to the WHO guidelines being 24% higher than that of their nulliparous peers[7]. However, not all mothers are similar.

In Denmark, almost 60.000 women give birth every year and out of those 20-22% give birth via caesarean section[8, 9], making caesarean section one of the most common surgical procedures in Denmark. Some studies suggest an association between caesarean section and a higher prevalence of postpartum depression or depressive symptoms compared with vaginal birth[10–13]. Regarding physical health issues, studies find that women who gave birth via caesarean section suffered more from exhaustion, lack of sleep, back pain, and bowel problems compared to women who gave birth vaginally[14–16]. As with all surgical operations, complications such as bleeding and infection might occur, and some women also experience chronic post-surgical pain following a caesarean section[17, 18]. Because of these psychological and physical differences between women who gave birth via caesarean section and women who gave birth vaginally, it is interesting to compare the two groups regarding levels of PA during the postpartum period, as potential differences in levels of PA may indicate a need for subgroup-specific interventions depending on mode of birth.

Therefore, we aimed to investigate whether there is a difference in non-adherence to the WHO guidelines of at least 150 minutes of moderate-to-vigorous physical activity (MVPA) per week within the first year postpartum, when comparing women who gave birth via caesarean section with women who gave birth vaginally. Based on the physical and psychological factors mentioned above, we hypothesise that postpartum women who gave birth via caesarean section adhere less to the WHO’s recommendations compared with women who gave birth vaginally.

## Methods

The reporting in this study follows STROBE guidelines (Appendix A).

### Study design

This population-based cross-sectional study was based on data from the Danish National Health Survey 2021[19] linked with data from the Danish Medical Birth Register[20] through the civil personal registration number, which is a unique 10-digit number given to all Danish citizens at birth[21].

Danish National Health Survey is a national survey conducted every four years to collect data on the population’s physical and mental health, health behaviour and morbidity[19]. The survey is distributed to a representative sample of the adult population (aged 16 years and above) from the five regions of Denmark, extracted from the Danish Civil Registration System. The survey is a self-administered questionnaire consisting of 56 questions[22, 23]. The design of the Danish National Health Survey is further described elsewhere[23]. The data in this study based on the 2021 Danish National Health Survey was collected from February 5 to May 12, 2021.

The Danish Medical Birth Register holds information on all births in Denmark including identification of parents by civil personal registration number, data on the birth including date, mode of birth, birth complications etc. and is maintained by the Danish Health Data Authority[20].

### Population

The study population was all Danish women aged 20-40 years, who completed the Danish National Health Survey in 2021 and gave birth to at least one live singleton child in the year prior to answering the Danish National Health Survey 2021. The childbirth status of each woman was found by linking to the Danish Medical Birth Register using their civil personal registration number[20–22].

### Exposure

The exposure variable was mode of birth. Data on this was obtained from the Danish Medical Birth Register[20]. The mothers were categorised into two groups based on diagnostic codes in the Danish Medical Birth Register: mothers who gave birth via caesarean section, which included all types of caesarean section, and mothers who had any form of vaginal birth (Appendix B).

### Outcome

The outcome was non-adherence to WHO guidelines, which was based on self-reported physical activity in the Danish National Health Survey. The amount of self-reported MVPA was based on the question: “In a typical week, how much time do you spend on moderate and vigorous physical activity in which you can feel a rise in breath” and self-reported vigorous PA was based on the question: “How much of the time, stated in the previous question, do you spend on vigorous physical activity, in which you can feel such a rise in breath that you are prevented from speaking?”. The answer categories contained five different options. For the MVPA question they were: (1) less than 30 minutes, (2) 30-89 minutes, (3) 90-149 minutes, (4) 150-299 minutes and (5) more than 300 minutes. For the vigorous PA question, they were: (1) less than 30 minutes, (2) 30-59 minutes, (3) 60-89 minutes, (4) 90-149 minutes and (5) more than 150 minutes. Based on these answers and the WHO guidelines of a minimum of 150 minutes of moderate PA, 75 minutes of vigorous PA or an equivalent combination of MVPA, the answers were dichotomised into mothers adhering to the WHO guidelines and mothers not adhering to the WHO guidelines as follows: If a person answered that they did at least 150 minutes of MVPA (answer 4 and 5) or at least 60 minutes of vigorous PA (answer 3, 4 and 5) per week they were categorised as adherent to the WHO guidelines. If they were below 150 minutes of MVPA (answer 1, 2 and 3) per week and 75 minutes of vigorous PA (answer 1 and 2) per week they were categorised as non-adherent to the WHO guidelines. If they did 90-149 minutes of MVPA (answer 3) per week and also did 30-59 minutes of vigorous PA (answer 2) per week they were categorised as adherent to the WHO guidelines.

### Demographics

Following demographics were extracted using the civil personal registration number and Danish administrative registries: Country of origin, employment status, cohabitation status, educational level, childbirth status, age, income in tertiles and urbanisation.

### Statistical analyses

Binomial regression was used to estimate the prevalence proportion ratio and the corresponding 95% confidence interval (CI) of mothers non-adhering to the WHO guidelines for PA, with mode of birth as the independent variable (vaginal birth as the reference group) and non-adherence as the dependent variable. Due to the estimand of interest being descriptive (i.e. the prevalence proportion ratio), no additional covariates were included[24]. Given that the dataset consists of all eligible women, no a priori sample size was calculated.

All statistical analyses were performed in R[25] with an alpha level of statistical significance set at 0.05.

## Results

The initial population for this study was women aged 20-40 years who answered the Danish National Health Survey in 2021 and who were registered in the newest version of the Danish Medical Birth Register, which holds data on births in Denmark from 2019 and onwards. A total of 3,773 mothers were included. Out of those, 87 were excluded because of multiple birth (not singleton) and 1,767 were excluded because of an age of their youngest child >1 year at the time they answered the survey. This left 1,919 postpartum mothers to be included in the main analysis (Figure 1).

**Figure 1:**
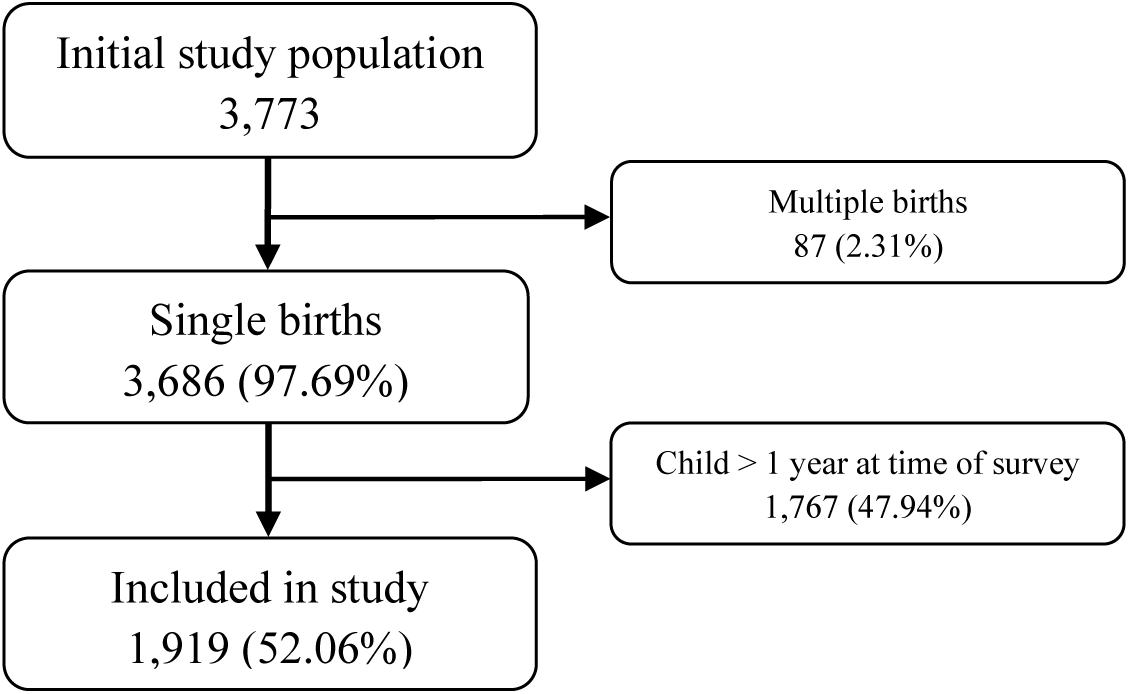
Flowchart of sampling process.

Descriptive characteristics of the study population is shown in table 1. Among the mothers in this study, 19.2% gave birth via caesarean section. Parity was split evenly in the group of mothers who gave birth via caesarean section, while 45.4% of mothers who gave birth vaginally was first-time mothers. Out of mothers who gave birth via caesarean section, 26% were in the age group 36-40 years, while that was only 16% for the women who gave birth vaginally.

**Table 1:** Characteristics of postpartum mothers included in the study (%)

|  | Vaginal birth | Caesarean section | Total |
| --- | --- | --- | --- |
|  | 1,550 (80.8) | 369 (19.2) | 1,919 (100) |
| **Country of origin** |  |  |  |
| Denmark | 1,143 (73.7) | 262 (71.0) | 1,405 (73.2) |
| Other western | 72 (4.6) | 19 (5.1) | 91 (4.7) |
| Non-western | 115 (7.4) | 34 (9.2) | 149 (7.8) |
| Unknown | 220 (14.2) | 54 (14.6) | 274 (14.3) |
| **Employment status** |  |  |  |
| Employed | 1,016 (65.5) | 255 (69.1) | 1,271 (66.2) |
| Unemployed<sup>a</sup> | 314 (20.3) | 60 (16.3) | 374 (19.5) |
| Unknown | 220 (14.2) | 54 (14.6) | 274 (14.3) |
| **Cohabitation<sup>b</sup>** |  |  |  |
| Living alone | 63 (4.1) | 12 (3.3) | 75 (3.9) |
| Living with someone | 1,267 (81.7) | 303 (82.1) | 1,570 (81.8) |
| Unknown | 220 (14.2) | 54 (14.6) | 274 (14.3) |
| **Educational level (years)** |  |  |  |
| ≤ 10 | 91 (5.9) | 24 (6.5) | 115 (6.0) |
| 11-15 | 437 (28.2) | 91 (24.7) | 528 (27.5) |
| ≥ 16 | 690 (44.5) | 159 (43.1) | 849 (44.2) |
| Unknown | 332 (21.4) | 95 (25.7) | 427 (22.3) |
| **Childbirth status** |  |  |  |
| Multipara | 847 (54.6) | 185 (50.1) | 1,032 (53.8) |
| First-time mothers | 703 (45.4) | 184 (49.9) | 887 (46.2) |
| **Age** |  |  |  |

**Table 1: Characteristics of postpartum mothers included in the study (%)**
|  | **Vaginal birth** | **Caesarean section** | **Total** |
| --- | --- | --- | --- |
|  | 1,550 (80.8) | 369 (19.2) | 1,919 (100) |
| 20-25 | 136 (8.8) | 23 (6.2) | 159 (8.3) |
| 26-30 | 586 (37.8) | 117 (31.7) | 703 (36.6) |
| 31-35 | 580 (37.4) | 133 (36.0) | 713 (37.2) |
| 36-40 | 248 (16.0) | 96 (26.0) | 344 (17.9) |
| **Income (tertiles)** |  |  |  |
| 1st | 245 (15.8) | 60 (16.3) | 305 (15.9) |
| 2nd | 507 (32.7) | 101 (27.4) | 608 (31.7) |
| 3rd | 577 (37.2) | 154 (41.7) | 731 (38.1) |
| Negative/Unknown | 221 (14.3) | 54 (14.6) | 275 (14.3) |
| **Urbanisation** |  |  |  |
| >100,000 | 479 (31.0) | 111 (30.1) | 590 (30.7) |
| 20,000-100,000 | 218 (14.1) | 48 (13.0) | 266 (13.9) |
| 1,000-20,000 | 367 (23.7) | 99 (26.8) | 466 (24.3) |
| <1,000 | 262 (16.9) | 57 (15.4) | 319 (16.6) |
| Unknown | 224 (14.5) | 54 (14.6) | 278 (14.5) |

<sup>a</sup>Including women enrolled in education or social security
<sup>b</sup>Based on civil status

The binomial regression showed a prevalence proportion of non-adherence to the WHO guidelines for PA of 70% (95% CI: [67; 72]%) among mothers who gave birth vaginally. When comparing mothers who gave birth via caesarean section with mothers who gave birth vaginally, we found a prevalence proportion ratio of 0.98 (95%-CI: [0.90; 1.06]), corresponding to an absolute difference in prevalence proportions of −1%-point (95%-CI: [−7; 4]%-point)

## Discussion

### Key findings

To our knowledge, no previous research has examined whether non-adherence to the WHO PA guidelines differs among postpartum women with different modes of birth in Denmark. In the present study, we found that there were no statistically significant differences in non-adherence to WHO PA guidelines between mothers who gave birth vaginally and mothers who gave birth via caesarean section.

This finding is in contrast to previous findings on other health outcomes following caesarean section[10, 13, 15, 16]. For instance, a meta-analysis from 2024 investigated the association between caesarean section and the risk of developing postpartum depression when comparing with mothers who had a vaginal birth[12]. They found an adjusted odds ratio for the association between caesarean section and postpartum depression within one to six months to be 1.29 (95% CI 1.18; 1.40), and after six months the adjusted odds ratio was 1.23 (95% CI 1.14; 1.33). Similarly, Woolhouse et al. found that women who gave birth via caesarean section were more likely to report extreme tiredness and back pain[14].

Based on these previous findings, we hypothesised, a priori, that postpartum women who gave birth via caesarean section adhere less to the WHO’s recommendations compared with postpartum women who gave birth vaginally. The lack of findings to support this hypothesis in the present study can have several explanations.

One potential explanation is that not all previous studies have shown a negative impact on health outcomes following caesarean section. For instance, Woolhouse et al. also found that women who gave birth vaginally suffered more from urinary incontinence than women who gave birth via caesarean section[14] and Spaich et al. reported that caesarean section did not influence the women’s sexual health[26]. Thus, our findings may support the notion that many factors and complex mechanisms may influence various health outcomes, such as physical activity, in the transition to motherhood[27, 28] and that the mode of birth has an intricate interplay with the postpartum period, which is a unique period in a woman’s life with many changes and new responsibilities.

Another potential explanation could be that the dichotomisation of PA into adherence or non-adherence to WHO PA guidelines, potentially masked any finer-grained differences in PA amount and/or intensity between the two groups of mothers. For instance, in a secondary analysis we saw that the proportion of mothers who reported more than 5 hours/week of moderate physical activity was statistically significantly higher among mothers who gave birth via caesarean section compared with mothers who gave birth vaginally (10.0% vs 5.4%; see Appendix C). This finding may suggest that mothers who give birth via caesarean section might be more polarised in their PA levels, with some of these mothers being very active while others are vastly less active.

A third potential explanation could be the time horizon for our analysis of one year postpartum, which were chosen because many physiological changes, such as cardiovascular remodelling, do not return to pre-pregnancy levels for at least one year after birth[29]. However, PA may normalise more rapidly than other physiological changes. A previous cohort study followed 471 women during pregnancy and postpartum regarding PA[30]. They found the levels of PA decreased from gestational week seventeen until birth and increased again at three months postpartum. Around one year postpartum the PA levels in the women had plateaued[30]. As such, we would expect any differences caused by the mode of birth to be most pronounced closer to the time of birth. However, in a secondary analysis (see appendix D) with a shorter time horizon of six months postpartum we did not see different findings, with a prevalence proportion ratio of 0.95 (95%-CI: [0.83; 1.06]). Other definitions of the postpartum period exist in the literature, such as the period a woman breastfeeds[31]. Ultimately, we chose the time horizon of one year postpartum for the main analysis both due to the aforementioned physiological changes and due to the fact that The Danish Health Authority recommends exclusive breastfeeding for the first six months postpartum and after this partially breastfeeding for at least twelve months or beyond[32].

### Limitations

One limitation to our study is that physical activity was self-reported through questionnaires. Even though the Danish National Health Survey was adjusted for non-response, it is possible that the self-reporting might have led to bias[22]. For instance, new mothers may have been overwhelmed, stressed out or suffer from a lack of sleep by their new role. If this led to different response rates from mothers who gave birth via caesarean section and mothers who gave birth vaginally this could lead to selection bias[14, 15]. Another limitation is that the Danish National Health Survey questionnaire provided only a couple of examples of MVPA, such as brisk walking, biking as transportation and garden work. These examples do not include examples of PA that mothers typically do, such as carrying the baby or walking with a stroller[33]. This might cause the mothers to underestimate their level of MVPA and thus under-report PA. On the other hand, people tend to overestimate abilities that are desirable in society such as PA, known as social desirability bias. For the biases mentioned in this section to impact the results of this study, it requires them to result in a difference in answers between mothers who gave birth via caesarean section and mothers who gave birth vaginally. Given that the factors above likely concern all mothers, we do not find it likely to have caused significant bias.

Another limitation that might influence the results of this study is the categorisation of births into either caesarean section or vaginal birth when there are different types of both caesarean sections and vaginal births. An American cohort study from 2022 looks at the risk of postpartum depression in different types of caesarean sections[13]. A total of 2,094 women were included, and they found a significant association between emergency caesarean section and postpartum depression with an adjusted odds ratio of 2.28 (95% CI 1.13; 4.57) compared to vaginal birth. They did not find a significant association between planned caesarean section and postpartum depression with an adjusted odds ratio of 1.96 (95% CI 0.72; 5.30)[13]. According to this study, there seems to be a difference in risk of postpartum depression between mothers who gave birth via a planned caesarean section and mothers who gave birth via an emergency caesarean section. We do not know if this could also reflect in their level of PA. It might dilute the estimate of mothers’ non-adherent to the WHO guidelines among mothers who had a caesarean section in the current study, when we have them all in the same subgroup no matter the type of caesarean section. Thus, we might have seen a difference in non-adherence if we had further differentiated caesarean section into emergency- and planned caesarean section.

A third limitation is that the 2021 Danish National Health Survey was collected during the COVID-19 pandemic, which caused change in the population’s behaviour, including habits of PA[34]. This has influenced the levels of PA in the entire population, and thus the difference in PA levels between the two groups of mothers is likely to be unaffected. However, the timing of the survey might limit the ability to compare non-adherence prevalence to other, non-pandemic points in time.

### Perspectives

Mothers tend to be less physical active than nulliparous women[35]. Danish mothers have a 24% higher non-adherence to the WHO guidelines compared with nulliparous women[7]. This highlights Danish mothers as a key population group for strategies and interventions aimed at promoting PA. To enable policy makers and healthcare professionals to allocate resources effectively and target the subgroups most in need, further stratification of Danish mothers is necessary to refine healthcare intervention. In this study we investigated whether mothers could be subgrouped based on mode of birth, as the Danish healthcare system currently does not differentiate postpartum healthcare interventions according to mode of birth. Based on the results of this study we do not find evidence to target interventions regarding PA based on mode of birth. However, mode of birth may be a relevant factor for targeting interventions for other health outcomes. This leaves future research to look for alternative ways to subgroup mothers, potentially identifying new subgroups to refine and better target intervention efforts. Nevertheless, greater attention to mothers is needed to support and promote higher levels of physical activity in this population of Danish women.

## Conclusion

This study was based on self-reported PA among 1,919 postpartum mothers responding to the Danish National Health Survey 2021 and linked with mode of birth in the Danish Medical Birth Register. Our hypothesis was that Danish mothers who gave birth via caesarean section had a higher prevalence proportion of non-adherence to the WHO PA guidelines of at least 150 minutes of MVPA per week, compared with mothers who gave birth vaginally. The statistical analysis showed no significant difference in non-adherence to the WHO PA guidelines between mothers who gave birth via caesarean section and mothers who gave birth vaginally with a PPR of 0.98 (95% CI 0.90; 1.06). The findings of this study do not suggest that future interventions, aiming at promoting PA among postpartum mothers in Denmark, should narrow the target group based on mode of birth. Future research may look for other possible identification of subgroups of Danish mothers to target interventions aimed at promoting PA in this population.

## Data Availability

All data produced in the present study are available upon reasonable request to the authors

## Acknowledgements

None

## References

1. Global status report on physical activity. 2022, World Health Organisation Geneva.

2. McMahon, E.M., et al., Physical activity in European adolescents and associations with anxiety, depression and well-being. Eur Child Adolesc Psychiatry, 2017. 26(1): p. 111–122.

3. Cheval, B., et al., Relationships between changes in self-reported physical activity, sedentary behaviour and health during the coronavirus (COVID-19) pandemic in France and Switzerland. J Sports Sci, 2021. 39(6): p. 699–704.

4. Mahindru, A., P. Patil, and V. Agrawal, Role of Physical Activity on Mental Health and Well-Being: A Review. Cureus, 2023. 15(1): p. e33475.

5. WHO guidelines on physical activity and sedentary behaviour. 2020, World Health Organization: Geneva.

6. Strain, T., et al., National, regional, and global trends in insufficient physical activity among adults from 2000 to 2022: a pooled analysis of 507 population-based surveys with 5&#xb7;7 million participants. The Lancet Global Health, 2024. 12(8): p. e1232–e1243.

7. Bueno, S.V., et al., Parous women perform less moderate to vigorous physical activity than their nulliparous peers: a population-based study in Denmark. Public Health, 2024. 231: p. 47–54.

8. Sundhedsstyrelsen, Anbefalinger for svangreomsorgen. 2022.

9. *Nyfødte og fødsler*. Available from: https://www.esundhed.dk/Emner/Graviditet-foedsler-og-boern/Nyfoedte-og-foedsler-1997-#tabpanel6B08359F7E54482C848E1F7625C740BD.

10. Xu, H., et al., Cesarean section and risk of postpartum depression: A meta-analysis. J Psychosom Res, 2017. 97: p. 118–126.

11. Orovou, E., et al., The Relation between Birth with Cesarean Section and Posttraumatic Stress in Postpartum Women. Maedica (Bucur), 2023. 18(4): p. 615–622.

12. Ning, J., et al., Meta-analysis of association between caesarean section and postpartum depression risk. Front Psychiatry, 2024. 15: p. 1361604.

13. Smithson, S., et al., Unplanned Cesarean delivery is associated with risk for postpartum depressive symptoms in the immediate postpartum period. J Matern Fetal Neonatal Med, 2022. 35(20): p. 3860–3866.

14. Woolhouse, H., et al., Physical health and recovery in the first 18 months postpartum: does cesarean section reduce long-term morbidity? Birth, 2012. 39(3): p. 221–9.

15. Thompson, J.F., et al., Prevalence and persistence of health problems after childbirth: associations with parity and method of birth. Birth, 2002. 29(2): p. 83–94.

16. Chen, H.H., et al., Understanding the relationship between cesarean birth and stress, anxiety, and depression after childbirth: A nationwide cohort study. Birth, 2017. 44(4): p. 369–376.

17. Jin, J., et al., Prevalence and risk factors for chronic pain following cesarean section: a prospective study. BMC Anesthesiol, 2016. 16(1): p. 99.

18. Weibel, S., et al., Incidence and severity of chronic pain after caesarean section: A systematic review with meta-analysis. Eur J Anaesthesiol, 2016. 33(11): p. 853–865.

19. Christensen, A.I., et al., 35 Years of health surveys in Denmark: a backbone of public health practice and research. Scand J Public Health, 2022. 50(7): p. 914–918.

20. Bliddal, M., et al., The Danish Medical Birth Register. Eur J Epidemiol, 2018. 33(1): p. 27–36.

21. Pedersen, C.B., The Danish Civil Registration System. Scand J Public Health, 2011. 39(7 Suppl): p. 22–5.

22. Christensen, A.I., et al., The Danish National Health Survey: Study design, response rate and respondent characteristics in 2010, 2013 and 2017. Scand J Public Health, 2022. 50(2): p. 180–188.

23. Sundhedsstyrelsen, *Danskernes Sundhed - Den Nationale Sundhedsprofil* 2021. 2021.

24. Lundberg, I., R. Johnson, and B.M. Stewart, What Is Your Estimand? Defining the Target Quantity Connects Statistical Evidence to Theory. American Sociological Review, 2021. 86(3): p. 532–565.

25. R Core Team (2021). R: A language and environment for statistical computing. R Foundation for Statistical Computing, Vienna, Austria. Version 4.4.1 URL https://www.R-project.org/

26. Spaich, S., et al., Influence of Peripartum Expectations, Mode of Delivery, and Perineal Injury on Women’s Postpartum Sexuality. J Sex Med, 2020. 17(7): p. 1312–1325.

27. ACOG Committee Opinion No. 736: Optimizing Postpartum Care. Obstet Gynecol, 2018. 131(5): p. e140–e150.

28. Finlayson, K., et al., What matters to women in the postnatal period: A meta-synthesis of qualitative studies. PLoS One, 2020. 15(4): p. e0231415.

29. Mottola, M.F., Exercise in the postpartum period: practical applications. Curr Sports Med Rep, 2002. 1(6): p. 362–8.

30. Borodulin, K., K.R. Evenson, and A.H. Herring, Physical activity patterns during pregnancy through postpartum. BMC Womens Health, 2009. 9: p. 32.

31. Evenson, K.R., et al., Summary of international guidelines for physical activity after pregnancy. Obstet Gynecol Surv, 2014. 69(7): p. 407–14.

32. Sundhedsstyrelsen. *Amning*. 25-06-2025]; Available from: https://www.sst.dk/da/Borger/En-sund-hverdag/Kost/Amning.

33. Adamo, K.B., et al., Young children and parental physical activity levels: findings from the Canadian health measures survey. Am J Prev Med, 2012. 43(2): p. 168–75.

34. Schmidt, T. and C.S. Pawlowski, Physical Activity in Crisis: The Impact of COVID-19 on Danes’ Physical Activity Behavior. Front Sports Act Living, 2020. 2: p. 610255.

35. Bellows-Riecken, K.H. and R.E. Rhodes, A birth of inactivity? A review of physical activity and parenthood. Prev Med, 2008. 46(2): p. 99–110.

