## Appendix A, B, C and D for "Mode of Birth and Adherence to WHO Physical Activity Guidelines: A Cross-Sectional Study of Danish Postpartum Mothers"

**Appendix A: STROBE statement**
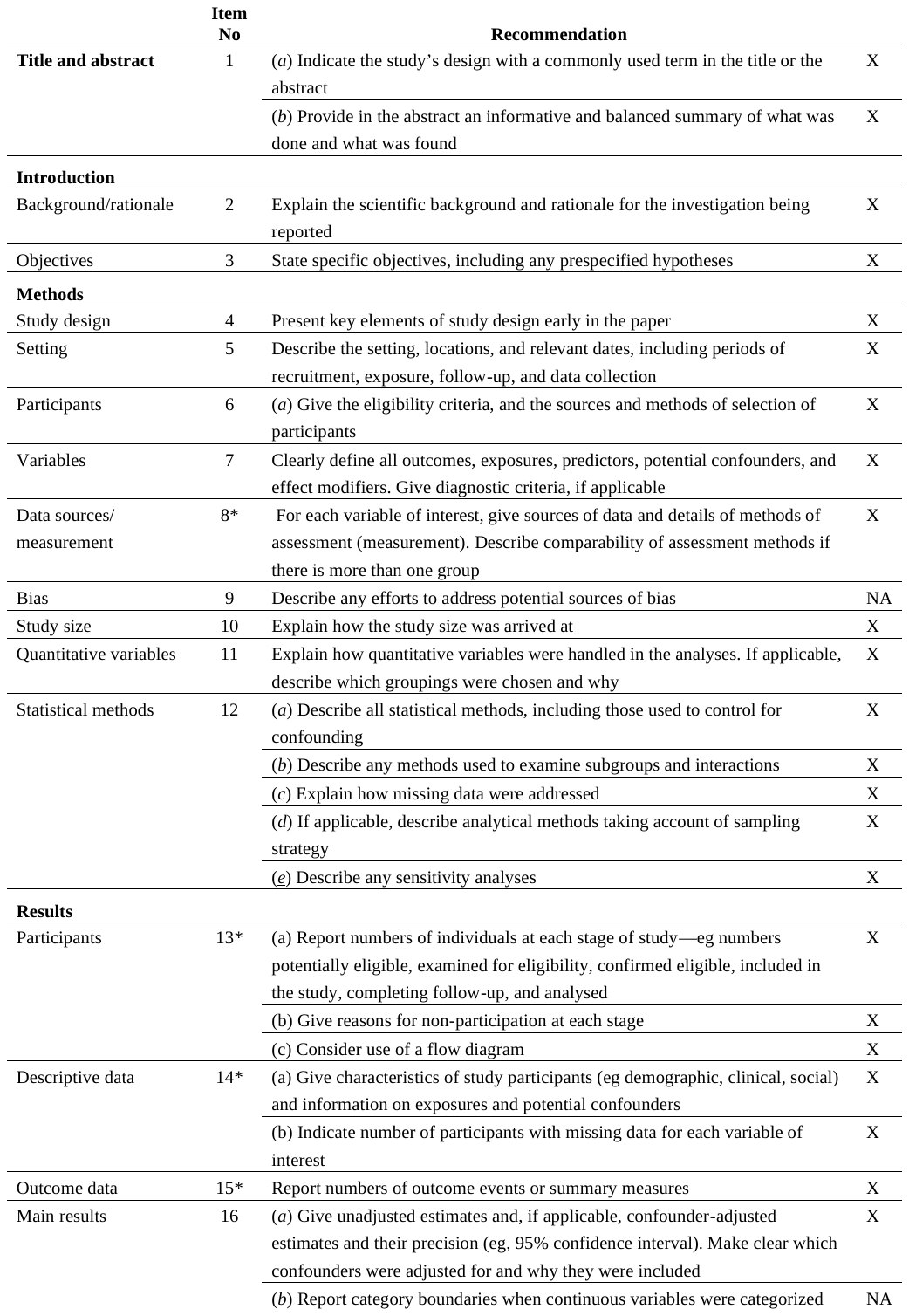

**Appendix B: Diagnostic codes used in the dataset**

DO809: Spontaneous singleton birth

DO819: Singleton birth with instrumental delivery

DO836: Singleton birth after abortion procedure

DO837: Singleton birth after induction

DO838: Other forms of singleton birth

DO829: Singleton birth via caesarean section

**Appendix C: MVPA by birth type**

### **Self-reported amount of moderate-to-vigorous physical activity (MVPA) by birth type as according to answers categories in the questionnaire. 95% CI = 95% confidence interval.**

|  | **Vaginal birth (N = 1550)** | | | Caesarean section (N = 369) | | |
| --- | --- | --- | --- | --- | --- | --- |
| Self-reported amount of MVPA | n | Proportion of group | 95% CI | **n** | **Proportion of group** | **95% CI** |
| <½ hour/week | 367 | 0.24 | (0.22, 0.26) | 102 | 0.28 | (0.23, 0.32) |
| ½–1½ hours/week | 431 | 0.28 | (0.26, 0.30) | 93 | 0.25 | (0.21, 0.30) |
| 1½–2½ hours/week | 237 | 0.15 | (0.13, 0.17) | 41 | 0.11 | (0.079, 0.14) |
| 2½–5 hours/week | 219 | 0.14 | (0.12, 0.16) | 47 | 0.13 | (0.093, 0.16) |
| >5 hours/week | 84 | 0.054 | (0.043, 0.065) | 37 | 0.10 | (0.070, 0.13) |
| Not reported | 212 | 0.14 | (0.12, 0.15) | 49 | 0.13 | (0.098, 0.17) |

**Appendix D: Sensitivity analysis for child age <=6 months**

Results from a sensitivity analysis similar to the main analysis except that only mothers with children aged 6 months or less are included. Women who gave birth via caesarean section did not report a different prevalence proportion of adhering to WHO guidelines regarding moderate-to-vigorous physical activity than women who gave birth vaginally (prevalence proportion ratio = 0.95, p = 0.362).

| Parameter | Prevalence proportion ratio | SE | 95% CI | z | p |
| --- | --- | --- | --- | --- | --- |
| (Intercept) | 0.70 | 0.02 | (0.66, 0.73) | -14.41 | <.001 |
| Caesarean section | 0.95 | 0.06 | (0.83, 1.06) | -0.91 | 0.362 |
